# BNT162b2 vaccine-induced humoral and cellular responses against SARS-CoV-2 variants in Systemic Lupus Erythematosus

**DOI:** 10.1101/2021.07.07.21260124

**Authors:** Quentin Moyon, Delphine Sterlin, Makoto Miyara, François Anna, Alexis Mathian, Raphael Lhote, Pascale Ghillani-Dalbin, Paul Breillat, Sasi Mudumba, Sophia de Alba, Fleur Cohen-Aubart, Julien Haroche, Micheline Pha, Thi Huong Du Boutin, Hedi Chaieb, Flores Pedro, Pierre Charneau, Guy Gorochov, Zahir Amoura

## Abstract

**Objectives:** Our aims were to evaluate Systemic Lupus Erythematosus (SLE) disease activity and SARS-CoV-2 specific immune responses after BNT162b2 vaccination.

**Methods:** In this prospective study, disease activity and clinical assessments were recorded from the first dose of vaccine, until day 15 after the second dose in 126 SLE patients. SARS-CoV-2 antibody responses were measured against wild-type spike antigen while serum-neutralizing activity was assessed against the SARS-CoV-2 historical strain and variants of concerns (VOCs). Vaccine-specific T-cell responses were quantified by Interferon (IFN)-γ release assay after the second dose.

**Results:** BNT162b2 was well tolerated and no statistically significant variations of BILAG and SLEDAI scores were observed throughout the study in SLE patients with active and inactive disease at baseline. Mycophenolate Mofetil (MMF) and Methotrexate (MTX) treatments were associated with drastically reduced BNT162b2 antibody-response (β=-78; p=0.007, β=-122; p<0.001, respectively). Anti-spike antibody response was positively associated with baseline total IgG serum levels, naïve B cell frequencies (β=2; p=0.018, β=2.5; p=0.003) and SARS-CoV-2-specific T cell response (r=0.462; p=0.003). In responders, serum neutralization activity decreased against VOCs bearing the E484K mutation but remained detectable in a majority of patients.

**Conclusion:** MMF, MTX and poor baseline humoral immune status, particularly: low naïve B cell frequencies, are independently associated with impaired BNT162b2 mRNA antibody response, delineating SLE patients who might need adapted vaccine regimens and follow-up.

**KEY MESSAGES:** *What is already known about this subject?:*

- BNT162b2 efficacy and safety has been described in studies mixing different RMDs

*What does this study add?:*

- No serious adverse effects, nor SLE flares have been documented after BNT162b2 in SLE patients.
- Not only MMF and MTX, but also a poor humoral immune status at baseline impair vaccine antibody response
- Albeit decreased, serum neutralizing activity against VOCs is conferred to vaccine-responders.

*How might this impact on clinical practice or future developments?:*

- These parameters could be helpful for physicians to delineate which patients should have antibody measurement after full BNT162b2 vaccination and should be proposed a third injection of BNT162b2 vaccine.

## INTRODUCTION

Because of the tremendous paucity of data on the impact of rheumatic and musculoskeletal diseases (RMD) and associated immune-modulatory treatments on Severe Acute Respiratory Coronavirus-2 (SARS-CoV-2) vaccination efficacy, most of the recommendations are currently based on expert opinions. Messenger RNA vaccination is a novel practice and its tolerance, immunogenicity and efficacy are poorly documented in RMD. Consequently, rules for vaccine against SARS-CoV2 vary according to country and over time [1,2]. Factors affecting the anti-SARS-CoV-2 antibody response have been explored, but only after a first dose or in studies mixing RMD [3,4]. Furthermore, the impact of treatments on the vaccine response is often studied mixing different RMDs [5]. Importantly, Simon *et al*. recently showed that inter-individual variations to vaccination were more related to the disease itself rather than to concomitant treatments [3]. Additionally, most of these studies focused on RMD treatments and not on the immunological status, which may also affect the antibody response. Among RMDs, SLE could represent a peculiar challenge to vaccination against SARS-CoV-2 [6]. The deregulation of type I interferon pathways associated with this condition [7] might impact on vaccine antibody response [8]. SLE-associated impaired lymphocyte functions might also impair vaccine efficacy [9,10]. Altogether, the risk of flares induced by vaccines is highly dependent on the disease studied and the specific scores used to measure this activity. It is therefore important to focus vaccine evaluation on homogeneous groups of patients.

Compared with the general population, SLE patients do not seem to be at higher risk of SARS-CoV-2 infections or severe COVID-19 [11–14], but this finding remains controversial as others studies found that SLE patients may be at higher risk of hospitalization during their COVID-19 course [15,16]. Increase of SLE disease activity has been previously reported during COVID-19 [17,18] but the risk of SLE flares following vaccination does not appears to be increased [18], although this point still requires confirmation through follow-up of SLE patients evaluated at identical pre- and post-vaccination time points in a prospective study. Finally, it remains unclear whether failures to induce antibody responses in patients under immunomodulatory regimens such as abatacept, mycophenolate mofetil (MMF), CD-20 inhibitors, calcineurin inhibitors[5,19] are also associated, or not, with an absence of vaccine-induced SARS-CoV-2-specific T cell responses. Here, we report post-vaccination disease activity data in 126 SLE patients, prospectively followed during the completion of a two-dose mRNA Pfizer/BioNTech (BNT162b2) vaccination regimen. SARS-CoV-2-specific humoral and cellular responses were monitored against the SARS-CoV-2 historical strain, but also against SARS-CoV-2 variants of concern (VOCs).

## METHODS

### Patients

The clinical study was conducted in the Internal Medicine Department 2, French National Reference Center for SLE, Pitié-Salpêtrière Hospital, Paris, France. Eligible patients were 18 years or older, with a diagnosis of SLE according to the revised American College of Rheumatology classification criteria [20]. Active lupus was defined with two scores: *(i)* at least 1 British Isles Lupus Assessment Group (BILAG) B in any organ, *(ii)* SLE Disease Activity Index (SLEDAI) 2K score > 4. Patients were vaccinated according to the French recommendations for Covid-19 vaccination [2]. The study protocol was approved by the Comité d’Ethique Sorbonne Université (CER-2021-011)

### Outcomes and follow-up

Patients were vaccinated at baseline (1st dose) against SARS-CoV-2 with Pfizer/BioNTech (BNT162b2) vaccine, and received the second dose at day (D)21-D28 unless contra-indicated. Patients were evaluated at baseline and at D7-14, D21-D28, D42. They were asked to contact their physician if they developed any symptoms in order to be promptly examined.

At each visit the following endpoints were assessed:

- Adverse events [21]
- SLE activity measured with SLEDAI 2K score [22,23] and BILAG score [24]
- SLE flares defined with the SELENA-SLEDAI flare index (SFI) [22,23] and BILAG 2004 score [24–26]
- SARS CoV-2 infection measured with anti-nucleocapsid antibodies
- Changes in serological activity (anti-dsDNA antibodies and C3), IFNα, anti-phospholipid antibodies [27]
- Anti-spike antibodies
- B, T and NK cells quantification
- B lymphocyte subsets

### Patient and public involvement

Patients were not involved in the design, or conduct, or reporting, or dissemination plans of this research.

### Serological analysis

SARS-CoV-2–specific IgG antibodies were measured as previously described [28]. Serum samples were tested with the Maverick SARSCoV-2 Multi-Antigen Serology Panel (Genalyte Inc., USA) according to the manufacturer’s instructions. The panel is designed to detect antibodies to five SARS-CoV-2 antigens: nucleocapsid, spike S1 Receptor Binding Domain (RBD), spike S1S2, spike S2, and spike S1, within a multiplex format based on photonic ring resonance technology. Briefly, 10 µl of each serum sample was added to a sample well plate array containing required diluents and buffers, and the plate and chip were loaded in the instrument for chip equilibration with the diluent buffer to measure baseline resonance. The serum sample was then charged over the chip to bind specific antibodies to antigens present on the chip. The chip was then washed to remove low-affinity binders, and specific antibodies were detected with anti-IgG secondary antibodies.

### Pseudoneutralization assay

Lentiviral particles carrying the luciferase gene and pseudotyped with spikes of SARS-CoV-2 historical strain or VOCs were produced by triple transfection of 293T cells as previously described [28]. Serum dilutions were mixed and co-incubated with 300 Transducing Units of pseudotyped lentiviral particles at room temperature for 30 min and then diluted in culture medium [Dulbecco’s modified Eagle’s medium–GlutaMAX (Gibco) + 10% fetal calf serum (Gibco) + 1% penicillin/streptomycin (Gibco)]. This mixture was then plated on tissue culture–treated black 96-well plates (Costar) with 20,000 HEK 293T-hACE2 cells per well in suspension. To prepare the suspension, cell flasks were washed with Dulbecco’s PBS (DPBS) twice (Gibco), and a single-cell suspension was made in DPBS + 0.1% EDTA (Promega) to preserve integrity of hACE2 protein. After 48 hours, the medium was removed from each well and bioluminescence was measured using a luciferase assay system (Promega) on an EnSpire plate reader (PerkinElmer).

### B cell phenotyping

B cell phenotyping was assessed on fresh whole blood. Briefly, 400μl of blood were washed in PBS1X-RPMI 5% (Gibco) then transferred in tubes containing anti-CD45 V500, anti-CD19 APC, anti-IgD FITC, anti-CD38 PerCPCy5.5, CD27 PE-Cy7, CD24 APC-H7, CD86 PE, CD3 BV421, CD14 BV421, CD21 BV421 lyophilized antibodies (BD Horizon™ Lyo technology). This lyophilized version of the multicolor panel increases the reagent stability and the assay performance. Cell staining was performed at room temperature for 15 minutes, then cells were washed and fixed (BD Cell Fix®). Cells were acquired on a BD FACS Canto II flow cytometer (BD Biosciences) and analyzed with FlowJo v10 software (FlowJo, LLC) according to the gating strategy presented in Figure S1.

### SARS-CoV-2-specific T cell responses

SARS-CoV-2-specific T cell responses were assessed in the clinical immunology laboratory of Pitié-Salpêtrière Hospital by a whole blood Interferon-Gamma Release Assay (IGRA) following manufacturer’s instructions (Quantiferon SARS-CoV-2, Qiagen). This test uses two Qiagen proprietary mixes of SARS-CoV-2 Spike protein (Ag.1 and Ag.2) selected to activate both CD4 and CD8 T cells. Briefly venous blood samples were transferred into the Quantiferon® tubes containing Spike peptides as well as positive and negative controls. Whole blood was incubated at 37°C for 16-24 hours and centrifuged to separate plasma. IFN-γ (IU/ml) was measured in these plasma samples using QuantiFERON Human IFN-γ SARS-CoV-2 ELISA kit (Qiagen) on Dynex DS2® analyzer (Qiagen).

### Statistical analysis

Baseline characteristics are reported with descriptive statistics. Linear regression models were used to assess the association between clinical and biological characteristics and the titer of IgG anti-RBD at day 42 in unadjusted and multivariable analysis. We considered potential confounders known or suspected to be associated with vaccine response such as demographic features (age, sex), activity of SLE, concomitant immune modulatory treatments and data from T, B and NK cells phenotyping. The beta coefficient is the degree of change in the outcome variable for every 1-unit of change in the predictor variable. If the beta coefficient is not statistically significant (*i*.*e*., the p-value is not significant), the variable does not significantly predict the outcome. If the beta coefficient is significant, examine the sign of the beta. If the beta coefficient is positive, the interpretation is that for every 1-unit increase in the predictor variable, the outcome variable will increase by the beta coefficient value. If the beta coefficient is negative, the interpretation is that for every 1-unit increase in the predictor variable, the outcome variable will decrease by the beta coefficient value. For example, if the beta coefficient is 0.80 and statistically significant, then for each 1-unit increase in the predictor variable, the outcome variable will increase by 0.80 units. Paired t-tests were used to detect differences in activity scores and biological data over time. As we excluded the 10 patients for whom follow-up was incomplete, we did not have to perform any imputation for missing data. Nonparametric test were used as Mann-Whitney U test to compare two independent groups, Wilcoxon test to compare paired values and Pearson coefficient to calculate correlation. Significant P values are indicated as described below: *p<0.05; **p<0.01; ***p<0.001; ****p<0.0001. Statistical analysis was performed using R software (version 4.1.0) and GraphPad Prism software, V6 (GraphPad, San Diego).

## RESULTS

### Demographic and disease characteristics

Vaccination against SARS-CoV-2 with Pfizer/BioNTech vaccine was proposed by their SLE referring physician to 180 patients with SLE; 127 (70.5 %) immediately accepted, 35 (19.4%) patients refused and 18 (10.0%) eventually accepted upon reflection, 9 of them were vaccinated in another center (Figure 1). A total of 136 SLE patients were enrolled and received one first dose; 3 patients received only one dose: either because they developed COVID-19 within 10 days after the first dose (n=2), or because COVID-19 had been contracted three months before the 1st dose (n=1). Among the 133 SLE patients who received 2 doses, 126 (92.6%) completed all the visits and were included in the final analysis. Baseline clinical characteristics of these 126 patients are summarized in Table 1. Treatments received from D1 to D42 were distributed as follows: hydroxychloroquine (n = 106; 84.1%; median daily dose : 400 mg), prednisone (n = 70 ; 55.5%) with 57 patients (45.2%) receiving less than 10mg daily (median daily dose : 5 mg) and 13 (10.3%) more than 10 mg daily (median daily dose : 19 mg), methotrexate (n = 20 ; 15.9% ; median weekly dose 15 mg) ; mycophenolate mofetil (n = 24 ; 19.0%; median daily dose = 2000 mg), azathioprine (n = 5 ; 4.0% ; median daily dose : 100 mg) and belimumab (n = 15 ; 11.9%), of whom 7 had intra-venous and 8 subcutaneous injections, respectively.

**Figure 1.**
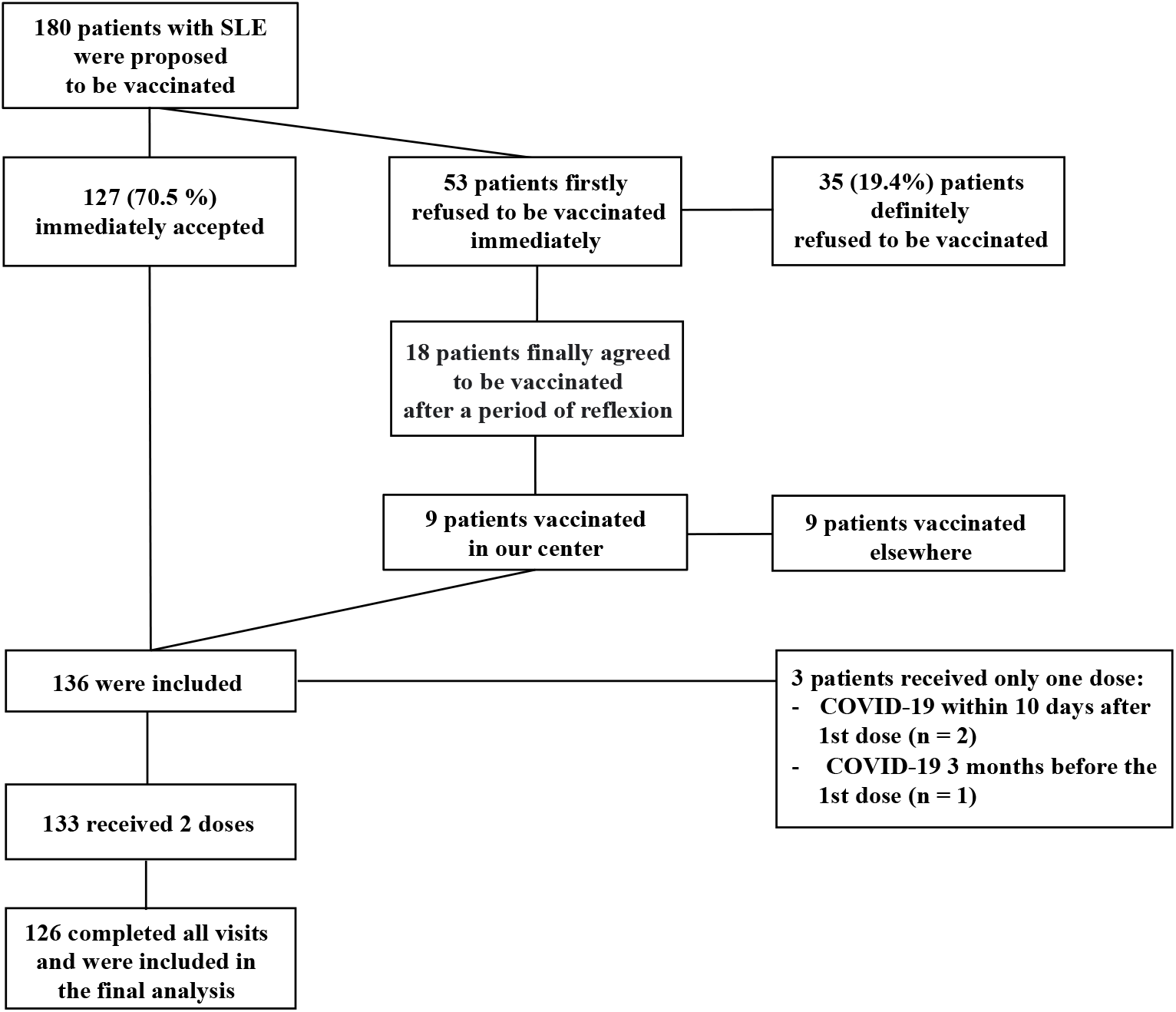
Study population and enrollment process. SLE patients were offered BNT162b2 vaccine through January 15, 2021.

**Table 1.**
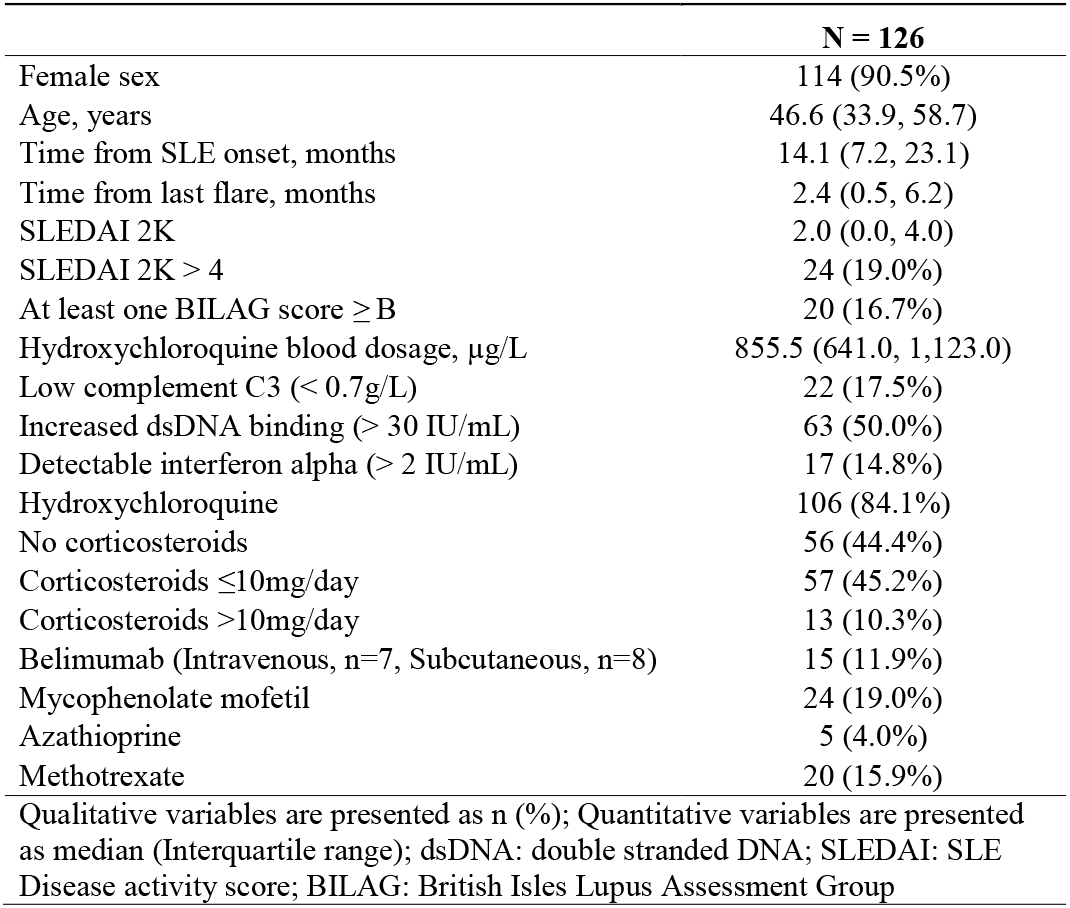
Demographics and clinico-biological features of SLE patients.

|  | **N = 126** |
| --- | --- |
| Female sex | 114 (90.5%) |
| Age, years | 46.6 (33.9, 58.7) |
| Time from SLE onset, months | 14.1 (7.2, 23.1) |
| Time from last flare, months | 2.4 (0.5, 6.2) |
| SLEDAI 2K | 2.0 (0.0, 4.0) |
| SLEDAI 2K > 4 | 24 (19.0%) |
| At least one BILAG score ≥ B | 20 (16.7%) |
| Hydroxychloroquine blood dosage, μg/L | 855.5 (641.0, 1,123.0) |
| Low complement C3 (< 0.7g/L) | 22 (17.5%) |
| Increased dsDNA binding (> 30 IU/mL) | 63 (50.0%) |
| Detectable interferon alpha (> 2 IU/mL) | 17 (14.8%) |
| Hydroxychloroquine | 106 (84.1%) |
| No corticosteroids | 56 (44.4%) |
| Corticosteroids ≤10mg/day | 57 (45.2%) |
| Corticosteroids >10mg/day | 13 (10.3%) |
| Belimumab (Intravenous, n=7, Subcutaneous, n=8) | 15 (11.9%) |
| Mycophenolate mofetil | 24 (19.0%) |
| Azathioprine | 5 (4.0%) |
| Methotrexate | 20 (15.9%) |
| Qualitative variables are presented as n (%); Quantitative variables are presented as median (Interquartile range); dsDNA: double stranded DNA; SLEDAI: SLE Disease activity score; BILAG: British Isles Lupus Assessment Group |  |

### Adverse BNT162b2 vaccine-associated events in SLE patients

No related serious adverse events (SAE), no grade 4 reactions, and no withdrawals due to related adverse events (AEs) were observed (Figure S2 and Table S1). Local reactions, predominantly pain at the injection site, were mild to moderate (grade 1 and 2).

### BNT162b2 vaccine effect on SLE disease activity

At baseline, 29 (23.0%) and 20 (16.7%) patients had active SLE according to SLEDAI (SLEDAI 2K > 4) and to BILAG (≥ 1 BILAG B), respectively. Within 42 days following vaccination (Figure 2A), mild disease flares were observed in 3 patients following vaccination, with a mucocutaneous BILAG score going from C to A in 1 individual, a musculoskeletal BILAG going from C to B in 1 individual and from D to C for 1 another vaccinated patient. In return, 9 patients (5 active and 4 inactive) clinically improved following vaccination with a musculoskeletal BILAG going from B to C for 4 patients and from C to D for 3 patients, a mucocutaneous BILAG going from A to C for 1 patient and a renal BILAG going from A to B for 1 patient. No statistically significant variation of SLEDAI score was observed throughout the study for active and inactive SLE patients according to initial SLEDAI score (SLEDAI 2K score ≤ 4 at day 1: mean[sd]; 1.2[1.4] day 1; 1.3[1.2] day 14; 1.0[1.2] day 28; 1.3[1.4] day 42, ns; SLEDAI score >4 at day 1: 11[5.1] day 1; 10.1[4.9] day 14; 10.0[5.3] day 28; 9.9[5.3] day 42, ns; Figure 2B). Altogether, vaccination is not preferentially associated with exacerbation of SLE symptoms, than with clinical improvement. When observed, variations of BILAG and SLEDAI scores were not preferentially observed in either active or inactive SLE patients at baseline.

**Figure 2.**
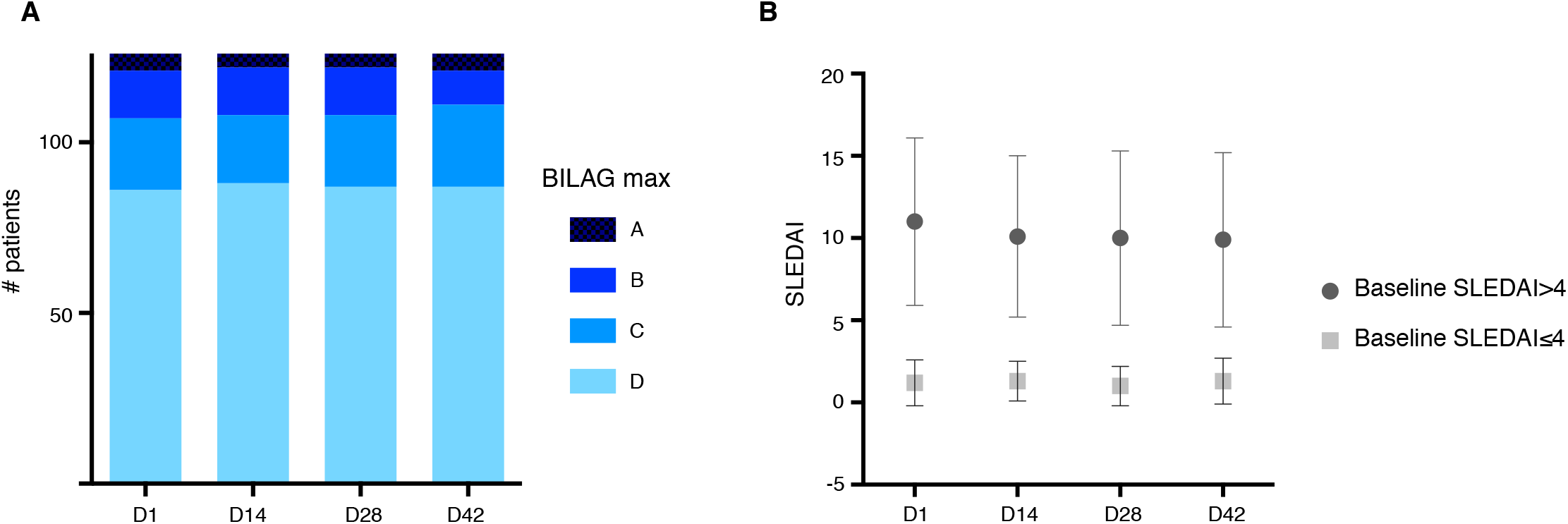
Evolution of SLE activity following vaccination. A. Repartition of maximal BILAG score at baseline and following vaccination B. Evolution of mean SLEDAI 2K score following vaccination

### Effect of treatments and baseline immune status on the immunogenicity of the BNT162b2 vaccine in SLE

Higher total serum IgG levels measured at baseline were associated with better seropositivity rates (β= 2.0; 95% CI: 0.34, 3.6; p = 0.018), while MMF and MTX uses were associated with lower anti-spike antibody production β = -78; 95% CI: -133, -22; p = 0.007 and β = -122; 95% CI: -184, -61; p <0.001, respectively) measured 14.7 days on average after the second injection (standard deviation 1.9 days). Total lymphocyte counts and IFNα levels at baseline were not significantly associated with seropositivity rates (Table 2). Hydroxychloroquine, steroids (either high or low dose) or belimumab use during the 42 days following vaccination did not impact anti-spike antibody production. Of note, SLE activity was not correlated with anti-spike antibody response, regardless of the score used to measure disease activity (see Table 2 and Table S2 with BILAG and SLEDAI, respectively).

**Table 2.** Baseline predictors of day 42 anti-SARS-CoV-2 RBD IgG titers according to linear regression model. SLE activity is measured with BILAG score.

| | $\beta$ | 95% CI | p-value |
| --- | --- | --- | --- |
| Age, years | -0.61 | -2.0, 0.75 | 0.4 |
| Male sex | -62 | -127, 3.7 | 0.064 |
| At least one BILAG score ≥ B | -45 | -106, 17 | 0.2 |
| C3, g/L | 35 | -77, 147 | 0.5 |
| dsDNA antibodies, IU/mL | 0.04 | -0.06, 0.13 | 0.4 |
| Detectable IFN alpha | -3.4 | -7.4, 0.58 | 0.093 |
| Total serum IgA, g/L | 1.8 | -12, 16 | 0.8 |
| Total serum IgG, g/L | 2.0 | 0.34, 3.6 | 0.018 |
| Total serum IgM, g/L | 12 | -1.0, 24 | 0.071 |
| Lymphocytes count, G/L | 6.6 | -31, 44 | 0.7 |
| Corticosteroids low | -20 | -66, 26 | 0.4 |
| Corticosteroids high | -50 | -127, 28 | 0.2 |
| Hydroxychloroquine | -27 | -85, 31 | 0.4 |
| Azathioprine | -118 | -242, 6.5 | 0.063 |
| Belimumab | -18 | -90, 54 | 0.6 |
| Mycophenolate mofetil | -78 | -133, -22 | 0.007 |
| Methotrexate | -122 | -184, -61 | <0.001 |
| Other immunosuppressor | 62 | -32, 156 | 0.2 |
| $\beta$: see Material and methods; BILAG: British Isles Lupus Assessment Group; CI = Confidence Interval; dsDNA: double stranded DNA; IFN: Interferon; RBD: Receptor Binding Domain | | | |

Since IgG levels but not total lymphocyte counts were significantly associated with the antibody response, we next studied the effect of lymphocyte sub-populations counts at baseline (Table 3).

**Table 3.** Baseline B, T and NK cells counts predictors of day 42 anti-SARS-CoV-2 RBD IgG titers according to linear regression model.

| | $\beta$ | 95% CI | p-value |
| --- | --- | --- | --- |
| B lymphocytes count, G/L | 0.38 | 0.13, 0.62 | 0.003 |
| NK lymphocytes count, G/L | 0.21 | -0.37, 0.80 | 0.5 |
| CD4+T lymphocytes count, G/L | 0.01 | -0.09, 0.11 | 0.9 |
| CD8+T lymphocytes, G/L | -0.01 | -0.16, 0.13 | 0.8 |
| $\beta$: see Material and methods; CI=Confidence Interval | | | |

We found that B lymphocyte counts were the sole lymphocyte population associated with anti-spike antibody response (β = 0.38; 95% CI: 0.13, 0.62; p=0.003). We further characterized the effect of B lymphocyte subsets at baseline. Treatments modifying B cell subpopulations were adjusted in this analysis (Table 4). Strikingly, naive B lymphocytes frequency at baseline was positively associated with anti-spike antibody response at D42 (β = 2.5; 95% CI; 0.87, 4.0; p=0.003; Table 4).

**Table 4.** Baseline B cell predictors of day 42 anti-SARS-CoV-2 RBD IgG titers according to linear regression model.

| | $\beta$ | 95% CI | p-value |
| --- | --- | --- | --- |
| Corticosteroids $\leq 10$mg/day | -42 | -93, 9.0 | 0.10 |
| Corticosteroids $> 10$mg/day | -135 | -269 | 0.007 |
| Hydroxychloroquine | -34 | -110, 43 | 0.4 |
| Azathioprine | -71 | -233, 92 | 0.4 |
| Belimumab | 9.6 | -105, 125 | 0.9 |
| Mycophenolate mofetil | -146 | -292 | $<0.001$ |
| Methotrexate | -121 | -243 | 0.004 |
| Other immunosuppressor | 203 | 5.6, 401 | 0.044 |
| Marginal zone B lymphocytes, Day 1 (%) | -0.22 | -2.7, 2.3 | 0.9 |
| Autoreactive B lymphocytes, Day 1 (%) | -1.5 | -4.3, 1.3 | 0.3 |
| Naive B Lymphocytes, Day 1 (%) | 2.5 | 0.87, 4.0 | 0.003 |
| Double negative B lymphocytes, Day 1 (%) | 3.8 | -2.2, 9.9 | 0.2 |
| Memory B lymphocytes, Day 1 (%) | -0.57 | -2.3, 1.2 | 0.5 |

$\beta$ : see Material and methods; CI=Confidence Interval; Autoreactive B cells (CD21lowCD38low); Double Negative B cells (CD27-IgD-); Marginal Zone B cells (CD27+IgD+); Memory B cells (CD27+IgD-); Naïve B cells (CD27-IgD+). B cell subsets frequencies are measured in total B cells.

### Effect of treatments on BNT162b2-induced neutralization responses

We next analyzed whether vaccine-induced antibody-responses may be protective by evaluating serum-neutralizing activity. As expected, we confirm a strong correlation between anti-RBD antibody levels and neutralization titers (SARS-CoV-2 D614G r=0.82, p<0.0001; Figure 3A). Consequently, parameters listed above influencing seroconversion also influenced neutralization activity (Table S3). MMF and MTX in particular have a negative impact on induction of neutralizing activity (β = -1.1; 95% CI: -1.9, -0.34; p = 0.005 and β = -1.9; 95% CI: -2.7; -1.1; p <0.001, respectively, Table S3). While a majority of MMF/MTX-treated patients still harbored detectable neutralizing activity (65% (15/23) MMF-treated patients, 68% (13/19) MTX-treated patients *vs* 96% (81/84) patients without MMF or MTX), their serum neutralizing activity drastically dropped compared to patients receiving other treatments (Inhibitory Dilution 50 (ID50) D614G median[min-max]; 111.2[30-18910] in MMF-treated patients *vs* 90.4[30-5527] in MTX-treated patients and 684.6[30-12061] in other patients; p<0.05; Figure 3B).

**Figure 3:**
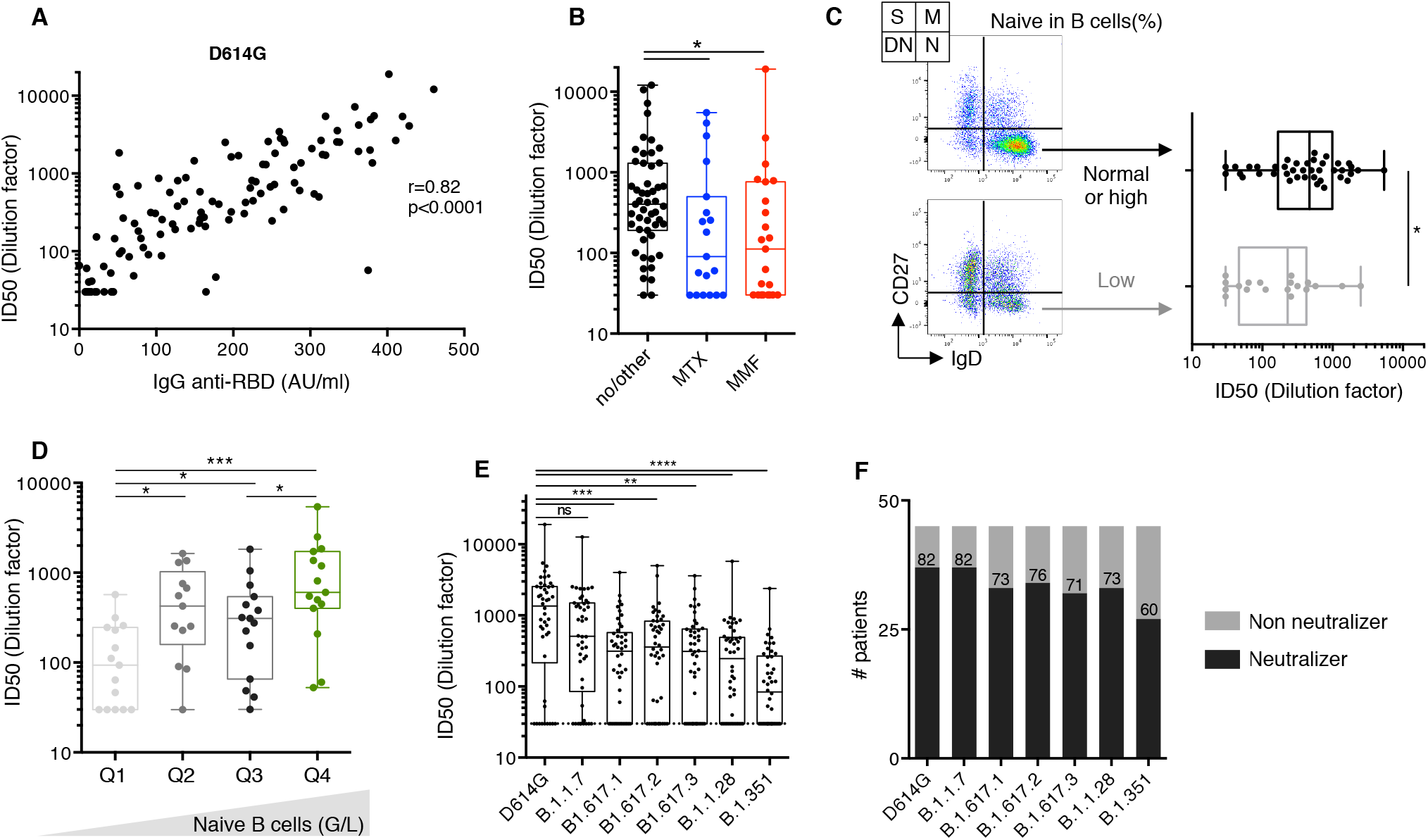
Vaccine-induced neutralizing potency. A. Comparison of serum anti-RBD IgG levels measured by photonic ring immunoassay with neutralizing capacity against D614G SARS-CoV-2 (n=126). Spearman coefficient (r) and p value (p) are indicated. B. Serum neutralizing activities against D614G SARS-CoV-2 measured as ID50 in 126 serum samples at D42. MTX- and MMF-treated patients are colour-coded (blue and red, respectively). Patients receiving other treatments are indicated in black. The boxplots show medians (middle line) and first and third quartiles while the whiskers indicate minimal and maximal values. P value was calculated using Kruskall-Wallis test (^*^ p<0.05). C. Comparison of serum neutralizing activities measured as ID50s against D614G SARS-CoV-2 in SLE patients with baseline low (grey, n=19) or high (black, n=40) naïve B cell frequency (arbitrary cut-off=42% of total B cells). Naïve B cells (N) are defined as CD27-Ig+ B cells, switched memory B cells (S) as CD27+IgD-, marginal zone B cells (M) as CD27+IgD+ and double negative B cells (DN) as CD27-IgD-. The boxplots show medians (middle line) and first and third quartiles while the whiskers indicate minimal and maximal values. P value was calculated using Mann-Whitney test (^*^ p<0.05). D. Serum neutralizing activities against D614G SARS-CoV-2 measured as ID50 in 59 SLE patients classified according to their naive B cells counts. Q1, Q2, Q3 and Q4 defined the naive B cell count quartiles. P value was calculated using Kruskall-Wallis test (^*^ p<0.05; ^***^ p<0.001). E. Serum neutralizing activities against indicated SARS-CoV-2 variants B.1.1.7 (Alpha), B.1.617.1 (Kappa), B.1.617.2 (Delta), B.1.617.3, B.1.28 (Gamma), and B.1.351 (Beta) measured as ID50 in 46 serum samples at D42. The boxplots show medians (middle line) and first and third quartiles while the whiskers indicate minimal and maximal values. P value was calculated using Kruskall-Wallis test (^**^ p<0.01; ^***^ p<0.001; ^****^ p<0.0001). F. Positive rates of serum neutralizing activity against SARS-CoV-2 variants in 46 SLE samples at day 42. Patient were defined as “neutralizers” (black) or “non-neutralizers” (grey) according to presence of neutralizing activity at first serum dilution (1/30), or not.

### Effect of baseline immune status on BNT162b2-induced neutralization responses

Consistent with serological studies, naive B cell decrease at baseline was negatively associated with serum D42 neutralizing activity (β = 0.04; 95% CI: 0.01, 0.07; p = 0.006; Table S4). As shown in Figure 3C, patients with a low naïve B cell compartment (<42% of B cells) developed a lower neutralizing activity than patients with normal or high naive B cell subset frequencies (229.2[30-2510] in low naïve B cell patients *vs* 468.3[30-5421]; p<0.05; Figure 3C). To more accurately evaluate the effect of naïve B cells on neutralizing antibody-response, we divided SLE patients in 4 groups according to their naïve baseline B cell counts (median[min-max] naïve B cell counts/μl: 9[0.01-23.2]; 41[27.2-50.9]; 68.1[57.2-98.7]; 133.8[110.1-160.2] in quartiles 1, 2, 3 and 4, respectively; Figure 3D). We confirm that naïve B cell counts are positively associated with vaccine-induced neutralizing antibody-responses (ID50 D614G 93.4[30-246.5] *vs* 340.1[30-1632] in quartiles 1 and 2, respectively; p<0.05, *vs* 315.2[30-721.1] in quartile 3 p<0.05; *vs* 679.9[60.4-2510] in quartile 4; p<0.001; Figure 3D).

These data therefore underline the importance of interrogating initial B cell status as well as immunosuppressive treatments to predict vaccine response.

### Broad neutralizing activity against VOCs in BNT162b2 vaccine responders

The Pfizer/BioNTech vaccine was designed to target the Wuhan isolate described by the end of 2019. However, emerging variants, with enhanced infectivity and the ability to escape immune control, rapidly became dominant. Concerns have been raised as to whether Pfizer/BioNTech vaccine will be effective against these emerging variants, particularly in vaccinated individuals receiving immunosuppressive drugs. We therefore measured neutralizing activities in the last 46 serum samples longitudinally collected against 4 major SARS-CoV-2 lineages: B.1.1.7 (originating in United Kingdom), B.1.351 (described in South Africa), B.1.1.28 (reported in Brazil) and B.1.617 (emerged in India). Consistent with previous studies [29,30], we found that vaccine-induced IgG antibodies efficiently cross-neutralize variants B.1.1.7 (Inhibitory Dilution 50 (ID50) median[min-max]; D614G 1453[30-18910] and B.1.1.7 514.5 [30-12625], ns, Figure 3E). It is noteworthy that serum neutralization activity decreased with lineages bearing the E484K mutation in the RBD (ID50 B1.617.1 341.1[30-3996], p<0.001; B.1.617.2 379.3[30-4982], p<0.001; B.1.617.3 317.9[30-3604], p<0.01; B.1.1.28 302.3[30-5757] and B.1.351 88.1[30-2389]; p<0.0001; Figure 3E), but remained detectable in a majority of patients (82% for B.1.1.7; 73% for B.1.617.1; 76% for B.1.617.2; 71% for B.1.617.3; 73% for B.1.1.28; 60% for B.1.351; Figure 3F). Among patients with neutralizing antibody activity against D614G strain, 100% (37/37) of patients also efficiently neutralized B.1.1.7 strain, 89% (33/37) B.1.617.1 variant, 92% (34/37) B.1.617.2 variant, 87% (32/37) B.1.1.28 variant, 89% (33/37) B.1.1.28 variant and 60% of patients (27/37) had detectable neutralizing activity against B.1.351.

Altogether these results demonstrated that vaccinated-SLE harbored decreased neutralizing activity against VOCs, as previously described in vaccinated healthy donors [31,32].

### SARS-CoV-2-specific T cell responses induced by the BNT162b2 vaccine in SLE

Beyond antibodies, T-cell immunity is required to confer optimal immune protection. In order to gain insight into the specific SARS-CoV-2 T cell response after vaccination in SLE patients, we evaluated IFN gamma secretion levels after specific T cell stimulation at day 15 after vaccination. While SARS-CoV-2-specific T cell responses were detected in 57% (17/30) of patients who had neutralizing antibody titers, cellular responses were only detected in 10% (1/10) of patients who had non-neutralizing antibody titers (p<0.05; Figure 4A). Interestingly, SARS-CoV-2-specific T cell responses were nevertheless detected in 2 out of 6 patients with very low levels of neutralizing activity in their serum (ID50 below 100 for D614G strain). Overall the strength of neutralizing antibody-response correlates with IFN-γ production by SARS-CoV-2 specific T cells (Antigen 1, r=0.462, p=0.003; Antigen 2 r=0.424, p=0.007, Figure 4B).

**Figure 4:**
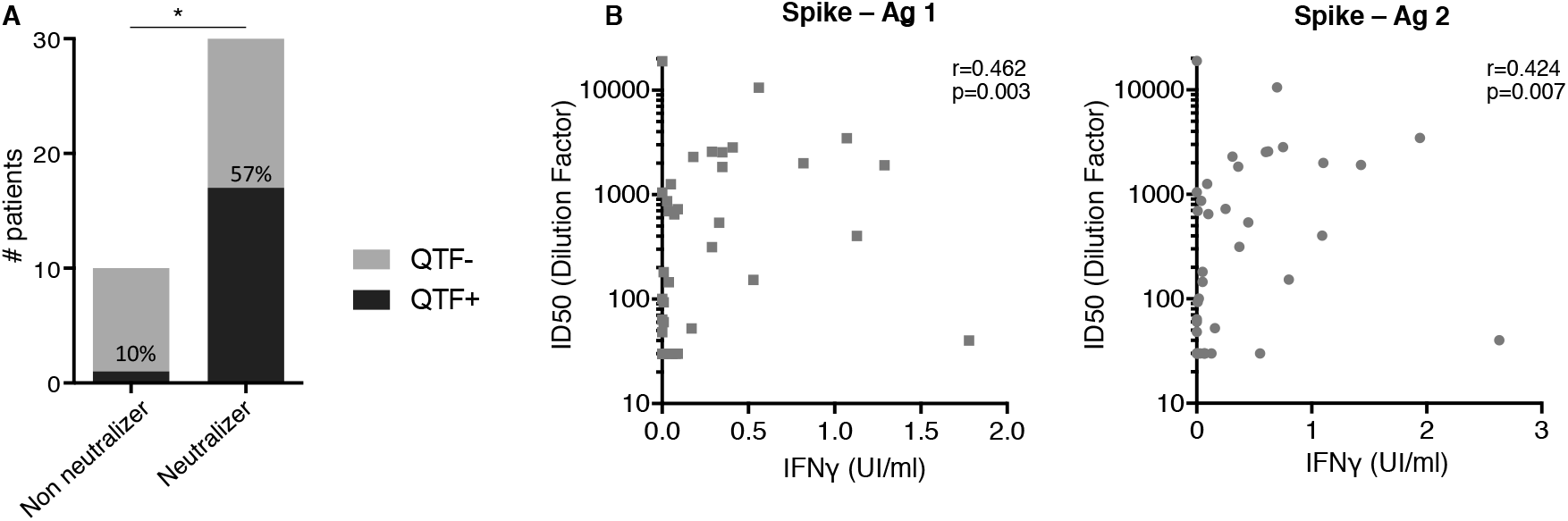
T-cell responses correlate with anti-SARS-CoV-2 humoral responses. A. Positives rates of Quantiferon SARS-CoV-2 testing in 40 SLE patients at D42, grouped according to serum neutralizer and non neutralizer status, as defined in Figure 3D. Numbers indicate the percentage of patients with a detectable T cell response. B. Comparison of IFNγ levels (UI/ml) after specific T cell stimulation using Quantiferon SARS-CoV-2 test and serum neutralizing activity reported with ID50 in 40 SLE patients at D45. Spearman coefficient (r) and p value (p) are indicated.

## DISCUSSION

Here, we report BNT162b2 antibody response measured both with anti-RBD antibody levels and neutralization activity in a cohort of 126 SLE french patients, with both active and inactive disease. To our knowledge, this is the first evaluation of BNT162b2-induced T cell and neutralization responses against VOCs in a cohort of SLE patients.

Global acceptance of BNT162b2 vaccine was 80.5%, in line with previous studies [33]. Most SLE patients were followed for a long time before vaccination in our center and vaccine was proposed by their treating physician. Interestingly, 18 (10%) patients who first refused vaccination, finally agreed to be vaccinated after a reflection time, a finding that is often lacking in Covid-19 vaccine acceptance studies. Tolerance of BNT162b2 vaccine was also good with a majority of local reactions and few systemic reactions.

SLE activity at time of vaccination, assessed either with the BILAG or the SLEDAI scores, neither reduced vaccine efficacy, nor increased the risk of subsequent SLE flares or vaccine side-effects. Consistent with this finding, previous meta-analysis of seasonal influenza and pneumococcal vaccinations in SLE demonstrated that immunization had no significant effect on the SLE activity measured with SLEDAI score [34]. Our results support the recommendation not to defer mRNA vaccination in active SLE patients [1]. One should note, however, that active SLE patients would subsequently receive treatments that could blunt BNT162b2 antibody-response. Indeed, MMF profoundly lowers BNT162b2 antibody-response as previously reported in transplant recipients [35] and RMDs patients [5]. MTX, a drug which is widely used for SLE, decreases Covid-vaccine antibody-response in a similar extent to MMF. Our results confirm recent studies [5,36] showing that MTX hampers immunogenicity to BNT162b2 mRNA Covid-19 vaccine in immune-mediated inflammatory diseases. However, since these two studies mixed different RMDs, the impact of these two drugs on BNT162b2 mRNA antibody response was assessed without adjusting with specific SLE parameters that could also affect BNT162b2 antibody-response (disease activity, IFNα levels). Reduced humoral responses to both seasonal influenza and pneumococcal vaccines with MTX in RA patients have been previously reported [37,38] while transitory MTX discontinuation improves the immunogenicity of seasonal influenza vaccination in RA patients [39–42]. Based on this RCT, the ACR recommended withholding MTX one week before each of the COVID-19 vaccine doses [1], but the evidence supporting this recommendation is unclear and was counterbalanced by the potential for RA flare associated with withholding MTX for a too long period, a recommendation that could not be extrapolated to SLE.

By contrast, neither hydroxychloroquine nor anti-BAFF belimumab did affect vaccine antibody-response. High-dose steroids were not associated either with a lower vaccine-induced antibody-response. The median prednisone daily high-dose was 19 mg in our study, a threshold that is lower than the one used for transplant recipient patients [43]. There is still controversy regarding the effect of steroids on SARS-CoV-2 vaccine efficacy, in particular whether a daily dose prednisone threshold above which antibody response might be blunted could be defined[1]. As a consequence, there is currently no expert panel recommendation to delay or not Covid vaccination in RMD patients receiving glucocorticoids at a prednisone-equivalent dose of ≥20 mg per day [1]. Optimal antibody responses seem to be elicited in RMDs patients on glucocorticoid monotherapy [44], although the daily prednisone dose was not reported in the latter study. Our data suggest that SLE patients with a daily dose of prednisone close to 20 mg should properly respond to BNT162b2 vaccine.

Elevated IFN-alpha serum levels were not associated with impaired BNT162b2 antibody-response, an observation in line with the lack of influence of SLE activity on vaccine efficacy. By contrast, elevated baseline total serum IgG levels were associated with a better antibody response. This association remains significant (p=0.018) when the analysis is adjusted for immunosuppressive drugs that could decrease IgG levels. For Chronic Lymphocytic Leukemia (CLL) patients, higher serum immunoglobulin levels at time of BNT162b2 mRNA vaccination were independently associated with a better response rate (IgG levels ≥ 550 mg/dL (OR 3.70, 95% CI 1.08-12.66)[45]. IgM levels were also an independently associated with serologic response (IgM≥ 40mg/dL (OR 2.92, 95% CI 1.21-7.02) in these patients. The influence of baseline IgG and IgM levels on COVID vaccine antibody-response have never been reported before in RMD and might be considered in future studies.

Our data also underline the importance of interrogating initial B cell compartments as correlates of predicted vaccine response. A marked decrease of naive B cells is known to be characteristic of SLE and not only the result of immunosuppressive drugs [46,47]. Here we observed a strong correlation of naive B cell loss with poor vaccine antibody-response, which likely points the role of naive B cells as a source of spike reactive-B cells. In recent studies extensive screening of pre-pandemic naive B cell repertoires revealed the presence of SARS-CoV-2-neutralizing antibody precursors. This subset of germline antibodies bound SARS-CoV-2 ACE2 receptor binding domain (RBD), albeit weakly, and may be engaged upon vaccine exposure to generate germinal centers and then follow affinity maturation process [48,49]. Indeed, Rincon-Arevalo *et al*. observed a significant difference in the frequency of SARS-CoV-2 RBD-specific naïve B cells between BNT162b2-responders and non-responders [50]. Reduced naive B cell pool in SLE would thus readily impact precursor frequency available to encounter the antigen, therefore impairing vaccine efficiency. Future vaccination strategies in SLE should consider naive B cells as an essential biomarker to define individuals at high risk of sub-optimal response that might benefit from reinforced vaccine regimens. It will remain to define in future studies whether patients eventually seroconverting after a third dose would have had readily detectable T cell responses after the second dose. Finally, much larger studies will be necessary to determine whether BNT162b2-induced T cell responses are solely sufficient to prevent at least from severe forms of the COVID-19 in patients.

Our study has some limitations. It is surprising to note that SARS-CoV-2-specific T cell responses were detected in only 57% of patients who had neutralizing antibody titers. This observation questions the sensitivity of the QTF assay used in our study and another [51]. Future studies should include other assays such T cell ELISPOT[52] to confirm this observation and whether low T cell responses would be more likely associated with SLE, compared to other RMDs and to healthy controls. Moreover, QTF assay was performed 15 days after the second dose, a timing that may be too short to optimally detect SARS-CoV-2 specific T cell response. Longitudinal studies are thus required to determine whether SLE patients develop a delayed cellular immune response. Unlike previous authors [3–5], we did not use antibody-response positivity thresholds. There are, however, no studies showing that these thresholds give RMD patients real protection against the risk of subsequent infection with SARS-CoV-2. It is not yet clear as to what immunogenicity parameter is predictive of vaccine-induced protection. Additionally, these thresholds vary according to the assays used and the variants studied, their clinical relevance is therefore questionable. To address this issue, Khoury *et al*. [53] recently analyzed the relationship between *in vitro* neutralization levels and the observed protection from SARS-CoV-2 infection using data from seven current vaccines and from convalescent cohorts. These authors found that despite expected inconsistencies between studies, comparison of normalized neutralization levels and vaccine efficacy demonstrates a remarkably strong non-linear relationship between mean neutralization level and the reported protection across different vaccines (Spearman r=0.905; P=0.0046). In this setting, the strong correlation we observed between RBD-antibody levels and the neutralizing activity is reassuring about the usefulness of serology in clinical practice. In our survey, only one patient out of 126 presented high IgG anti-RBD levels and low neutralizing activity (Figure 3A). Antibody response was assessed 14 days after the second injection. We cannot rule out the hypothesis that a higher antibody response would have been observed later [44]. Of note, Polack *et al*. measured antibody responses as soon as 7 days after second injection [21] and were able to link BNT162b2 efficacy to prevention of SARS-CoV-2 infections in healthy individuals. Last, this SLE cohort did not comprise rituximab-treated patients, in whom antibody responses are abrogated [54]. Rituximab is not approved for SLE although used in clinical practice.

Despite its limitations, this study provides evidence that in SLE, use of MMF or MTX is associated with reduced vaccine efficacy. We also show that low baseline IgG levels and a reduced pool of naive B cells are predictive of impaired vaccination-induced neutralizing activity against SARS-CoV-2. These parameters could be helpful for physicians to delineate which patients should have antibody measurement after full BNT162b2 vaccination and should be proposed a third injection of BNT162b2 vaccine.

## Data Availability

All data are available in the manuscript or the supplementary materials.

## ACKNOWLEDGMENTS

The authors wish to thank the patients who agreed to participate in this study, nurses from the Internal Medicine Department of Institut E3M (Assistance Publique Hôpitaux de Paris (AP-HP), Hôpital Pitié-Salpêtrière, 75013 Paris, France) who participated in this study, all members from the Immunology Department, (Assistance Publique Hôpitaux de Paris (AP-HP), Hôpital Pitié-Salpêtrière, 75013 Paris, France), who performed Quantiferon analysis and B-cell phenotyping, Laura Wakselman from clinical research unit (URC) of Pitié-Salpêtrière Hospital for help with regulatory and ethical issues.

## CONTRIBUTORSHIP

Q.M, A.M, R.L, F.C-A, J.H, M.P, L.B, Z.A recruited patients. Q.M, P.B, C.H, F.P, P.G collected demographic and clinico-biological data. M.M, D.S, P.G prepared the specimens. M.M, Q.M, F.A, D.S designed and performed experiments. Q.M, D.S, F.A, M.M, S.A, S.M analyzed data. Q.M, D.S prepared the figures. Q.M, D.S, G.G, Z.A wrote the manuscript. Q.M, D.S, F.A, M.M, G.G, Z.A designed the study and reviewed the manuscript.

## DATA SHARING STATEMENT

All data are available in the manuscript or the supplementary materials.

## ETHICAL APPROVAL INFORMATION

The study protocol was approved by the Comité d’Ethique Sorbonne Université (CER-2021-011)

## REFERENCES

1 Curtis JR, Johnson SR, Anthony DD, et al. American College of Rheumatology Guidance for COVID-19 Vaccination in Patients With Rheumatic and Musculoskeletal Diseases: Version 1. Arthritis & Rheumatology 2021;n/a. doi:10.1002/art.41734

2 Haut Conseil de la santé publique. Avis complémentaire à l’avis du 14 janvier relatif aux mesures de contrôle et de prévention de la diffusion des nouveaux variants du SARS-CoV-2. 2021.

3 Simon D, Tascilar K, Fagni F, et al. SARS-CoV-2 vaccination responses in untreated, conventionally treated and anticytokine-treated patients with immune-mediated inflammatory diseases. Ann Rheum Dis Published Online First: 6 May 2021. doi:10.1136/annrheumdis-2021-220461

4 Boyarsky BJ, Ruddy JA, Connolly CM, et al. Antibody response to a single dose of SARS-CoV-2 mRNA vaccine in patients with rheumatic and musculoskeletal diseases. Ann Rheum Dis Published Online First: 23 March 2021. doi:10.1136/annrheumdis-2021-220289

5 Furer V, Eviatar T, Zisman D, et al. Immunogenicity and safety of the BNT162b2 mRNA COVID-19 vaccine in adult patients with autoimmune inflammatory rheumatic diseases and in the general population: a multicentre study. Ann Rheum Dis Published Online First: 14 June 2021. doi:10.1136/annrheumdis-2021-220647

6 Tang W, Askanase AD, Khalili L, et al. SARS-CoV-2 vaccines in patients with SLE. Lupus Sci Med 2021;8. doi:10.1136/lupus-2021-000479

7 Postal M, Vivaldo JF, Fernandez-Ruiz R, et al. Type I interferon in the pathogenesis of systemic lupus erythematosus. Curr Opin Immunol 2020;67:87–94. doi:10.1016/j.coi.2020.10.014

8 Björk A, Da Silva Rodrigues R, Richardsdotter Andersson E, et al. Interferon activation status underlies higher antibody response to viral antigens in patients with systemic lupus erythematosus receiving no or light treatment. Rheumatology (Oxford) Published Online First: 2 October 2020. doi:10.1093/rheumatology/keaa611

9 Chen P-M, Tsokos GC. T Cell Abnormalities in the Pathogenesis of Systemic Lupus Erythematosus: an Update. Curr Rheumatol Rep 2021;23:12. doi:10.1007/s11926-020-00978-5

10 Mathian A, Devilliers H, Krivine A, et al. Factors influencing the efficacy of two injections of a pandemic 2009 influenza A (H1N1) nonadjuvanted vaccine in systemic lupus erythematosus. Arthritis Rheum 2011;63:3502–11. doi:10.1002/art.30576

11 Mathian A, Mahevas M, Rohmer J, et al. Clinical course of coronavirus disease 2019 (COVID-19) in a series of 17 patients with systemic lupus erythematosus under long-term treatment with hydroxychloroquine. Ann Rheum Dis 2020;79:837–9. doi:10.1136/annrheumdis-2020-217566

12 Ramirez GA, Gerosa M, Beretta L, et al. COVID-19 in systemic lupus erythematosus: Data from a survey on 417 patients. Semin Arthritis Rheum 2020;50:1150–7. doi:10.1016/j.semarthrit.2020.06.012

13 Favalli EG, Gerosa M, Murgo A, et al. Are patients with systemic lupus erythematosus at increased risk for COVID-19? Ann Rheum Dis 2021;80:e25. doi:10.1136/annrheumdis-2020-217787

14 Gartshteyn Y, Askanase AD, Schmidt NM, et al. COVID-19 and systemic lupus erythematosus: a case series. Lancet Rheumatol 2020;2:e452–4. doi:10.1016/S2665-9913(20)30161-2

15 Fernandez-Ruiz R, Masson M, Kim MY, et al. Leveraging the United States Epicenter to Provide Insights on COVID-19 in Patients With Systemic Lupus Erythematosus. Arthritis Rheumatol 2020;72:1971–80. doi:10.1002/art.41450

16 Gianfrancesco M, Hyrich KL, Al-Adely S, et al. Characteristics associated with hospitalisation for COVID-19 in people with rheumatic disease: data from the COVID-19 Global Rheumatology Alliance physician-reported registry. Ann Rheum Dis 2020;79:859–66. doi:10.1136/annrheumdis-2020-217871

17 Raghavan S, Gonakoti S, Asemota IR, et al. A Case of Systemic Lupus Erythematosus Flare Triggered by Severe Coronavirus Disease 2019. J Clin Rheumatol 2020;26:234–5. doi:10.1097/RHU.0000000000001531

18 Kondo Y, Kaneko Y, Oshige T, et al. Exacerbation of immune thrombocytopaenia triggered by COVID-19 in patients with systemic lupus erythematosus. Ann Rheum Dis Published Online First: 5 August 2020. doi:10.1136/annrheumdis-2020-218157

19 Rozen-Zvi B, Yahav D, Agur T, et al. Antibody response to SARS-CoV-2 mRNA vaccine among kidney transplant recipients: a prospective cohort study. Clin Microbiol Infect Published Online First: 3 May 2021. doi:10.1016/j.cmi.2021.04.028

20 Hochberg MC. Updating the American College of Rheumatology revised criteria for the classification of systemic lupus erythematosus. Arthritis Rheum 1997;40:1725. doi:10.1002/art.1780400928

21 Polack FP, Thomas SJ, Kitchin N, et al. Safety and Efficacy of the BNT162b2 mRNA Covid-19 Vaccine. N Engl J Med 2020;383:2603–15. doi:10.1056/NEJMoa2034577

22 Buyon JP, Petri MA, Kim MY, et al. The effect of combined estrogen and progesterone hormone replacement therapy on disease activity in systemic lupus erythematosus: a randomized trial. Ann Intern Med 2005;142:953–62. doi:10.7326/0003-4819-142-12_part_1-200506210-00004

23 Petri M, Kim MY, Kalunian KC, et al. Combined oral contraceptives in women with systemic lupus erythematosus. N Engl J Med 2005;353:2550–8. doi:10.1056/NEJMoa051135

24 Isenberg DA, Rahman A, Allen E, et al. BILAG 2004. Development and initial validation of an updated version of the British Isles Lupus Assessment Group’s disease activity index for patients with systemic lupus erythematosus. Rheumatology (Oxford) 2005;44:902–6. doi:10.1093/rheumatology/keh624

25 Gordon C, Sutcliffe N, Skan J, et al. Definition and treatment of lupus flares measured by the BILAG index. Rheumatology (Oxford) 2003;42:1372–9. doi:10.1093/rheumatology/keg382

26 Isenberg DA, Allen E, Farewell V, et al. An assessment of disease flare in patients with systemic lupus erythematosus: a comparison of BILAG 2004 and the flare version of SELENA. Ann Rheum Dis 2011;70:54–9. doi:10.1136/ard.2010.132068

27 Mathian A, Mouries-Martin S, Dorgham K, et al. Monitoring Disease Activity in Systemic Lupus Erythematosus With Single-Molecule Array Digital Enzyme-Linked Immunosorbent Assay Quantification of Serum Interferon-α. Arthritis Rheumatol 2019;71:756–65. doi:10.1002/art.40792

28 Sterlin D, Mathian A, Miyara M, et al. IgA dominates the early neutralizing antibody response to SARS-CoV-2. Sci Transl Med 2021;13. doi:10.1126/scitranslmed.abd2223

29 Sahin U, Muik A, Derhovanessian E, et al. COVID-19 vaccine BNT162b1 elicits human antibody and T(H)1 T cell responses. Nature 2020;586:594–9. doi:10.1038/s41586-020-2814-7

30 Liu J, Liu Y, Xia H, et al. BNT162b2-elicited neutralization of B.1.617 and other SARS-CoV-2 variants. Nature Published Online First: 10 June 2021. doi:10.1038/s41586-021-03693-y

31 Planas D, Veyer D, Baidaliuk A, et al. Reduced sensitivity of infectious SARS-CoV-2 variant B.1.617.2 to monoclonal antibodies and sera from convalescent and vaccinated individuals. bioRxiv 2021;:2021.05.26.445838. doi:10.1101/2021.05.26.445838

32 Ferreira I, Datir R, Papa G, et al. SARS-CoV-2 B.1.617 emergence and sensitivity to vaccine-elicited antibodies. bioRxiv 2021;:2021.05.08.443253. doi:10.1101/2021.05.08.443253

33 Campochiaro C, Trignani G, Tomelleri A, et al. Potential acceptance of COVID-19 vaccine in rheumatological patients: a monocentric comparative survey. Ann Rheum Dis Published Online First: 28 January 2021. doi:10.1136/annrheumdis-2020-219811

34 Pugès M, Biscay P, Barnetche T, et al. Immunogenicity and impact on disease activity of influenza and pneumococcal vaccines in systemic lupus erythematosus: a systematic literature review and meta-analysis. Rheumatology (Oxford) 2016;55:1664–72. doi:10.1093/rheumatology/kew211

35 Boyarsky BJ, Werbel WA, Avery RK, et al. Immunogenicity of a Single Dose of SARS-CoV-2 Messenger RNA Vaccine in Solid Organ Transplant Recipients. JAMA 2021;325:1784–6. doi:10.1001/jama.2021.4385

36 Haberman RH, Herati R, Simon D, et al. Methotrexate hampers immunogenicity to BNT162b2 mRNA COVID-19 vaccine in immune-mediated inflammatory disease. Ann Rheum Dis Published Online First: 25 May 2021. doi:10.1136/annrheumdis-2021-220597

37 Kapetanovic MC, Nagel J, Nordström I, et al. Methotrexate reduces vaccine-specific immunoglobulin levels but not numbers of circulating antibody-producing B cells in rheumatoid arthritis after vaccination with a conjugate pneumococcal vaccine. Vaccine 2017;35:903–8. doi:10.1016/j.vaccine.2016.12.068

38 Kapetanovic MC, Roseman C, Jönsson G, et al. Antibody response is reduced following vaccination with 7-valent conjugate pneumococcal vaccine in adult methotrexate-treated patients with established arthritis, but not those treated with tumor necrosis factor inhibitors. Arthritis Rheum 2011;63:3723–32. doi:10.1002/art.30580

39 Park JK, Lee MA, Lee EY, et al. Effect of methotrexate discontinuation on efficacy of seasonal influenza vaccination in patients with rheumatoid arthritis: a randomised clinical trial. Ann Rheum Dis 2017;76:1559–65. doi:10.1136/annrheumdis-2017-211128

40 Park JK, Lee YJ, Shin K, et al. Impact of temporary methotrexate discontinuation for 2 weeks on immunogenicity of seasonal influenza vaccination in patients with rheumatoid arthritis: a randomised clinical trial. Ann Rheum Dis 2018;77:898–904. doi:10.1136/annrheumdis-2018-213222

41 Park JK, Kim MJ, Choi Y, et al. Effect of short-term methotrexate discontinuation on rheumatoid arthritis disease activity: post-hoc analysis of two randomized trials. Clin Rheumatol 2020;39:375–9. doi:10.1007/s10067-019-04857-y

42 Park JK, Choi Y, Winthrop KL, et al. Optimal time between the last methotrexate administration and seasonal influenza vaccination in rheumatoid arthritis: post hoc analysis of a randomised clinical trial. Ann Rheum Dis 2019;78:1283–4. doi:10.1136/annrheumdis-2019-215187

43 Rabinowich L, Grupper A, Baruch R, et al. Low immunogenicity to SARS-CoV-2 vaccination among liver transplant recipients. J Hepatol Published Online First: 21 April 2021. doi:10.1016/j.jhep.2021.04.020

44 Ruddy JA, Connolly CM, Boyarsky BJ, et al. High antibody response to two-dose SARS-CoV-2 messenger RNA vaccination in patients with rheumatic and musculoskeletal diseases. Ann Rheum Dis Published Online First: 24 May 2021. doi:10.1136/annrheumdis-2021-220656

45 Herishanu Y, Avivi I, Aharon A, et al. Efficacy of the BNT162b2 mRNA COVID-19 vaccine in patients with chronic lymphocytic leukemia. Blood 2021;137:3165–73. doi:10.1182/blood.2021011568

46 Odendahl M, Jacobi A, Hansen A, et al. Disturbed peripheral B lymphocyte homeostasis in systemic lupus erythematosus. J Immunol 2000;165:5970–9. doi:10.4049/jimmunol.165.10.5970

47 Dörner T, Jacobi AM, Lee J, et al. Abnormalities of B cell subsets in patients with systemic lupus erythematosus. J Immunol Methods 2011;363:187–97. doi:10.1016/j.jim.2010.06.009

48 Kreer C, Zehner M, Weber T, et al. Longitudinal Isolation of Potent Near-Germline SARS-CoV-2-Neutralizing Antibodies from COVID-19 Patients. Cell 2020;182:843-854.e12. doi:10.1016/j.cell.2020.06.044

49 Feldman J, Bals J, Denis KSt, et al. Naive human B cells can neutralize SARS-CoV-2 through recognition of its receptor binding domain. bioRxiv 2021;:2021.02.02.429458. doi:10.1101/2021.02.02.429458

50 Rincon-Arevalo H, Choi M, Stefanski A-L, et al. Impaired humoral immunity to SARS-CoV-2 BNT162b2 vaccine in kidney transplant recipients and dialysis patients. Sci Immunol 2021;6. doi:10.1126/sciimmunol.abj1031

51 Van Praet JT, Vandecasteele S, De Roo A, et al. Humoral and cellular immunogenicity of the BNT162b2 mRNA Covid-19 Vaccine in nursing home residents. Clin Infect Dis Published Online First: 7 April 2021. doi:10.1093/cid/ciab300

52 Zuo J, Dowell AC, Pearce H, et al. Robust SARS-CoV-2-specific T cell immunity is maintained at 6 months following primary infection. Nat Immunol 2021;22:620–6. doi:10.1038/s41590-021-00902-8

53 Khoury DS, Cromer D, Reynaldi A, et al. Neutralizing antibody levels are highly predictive of immune protection from symptomatic SARS-CoV-2 infection. Nat Med Published Online First: 17 May 2021. doi:10.1038/s41591-021-01377-8

54 D’Silva KM, Serling-Boyd N, Hsu TY-T, et al. SARS-CoV-2 antibody response after COVID-19 in patients with rheumatic disease. Ann Rheum Dis Published Online First: 12 January 2021. doi:10.1136/annrheumdis-2020-219808

