## Supplemental material for "BNT162b2 vaccine-induced humoral and cellular responses against SARS-CoV-2 variants in Systemic Lupus Erythematosus"

**SUPPLEMENTARY MATERIAL**

**Table S1: Local and systemic adverse events reported at days 14, 28 and 45 after first BNT162b2 dose in 126 SLE patients**

| Adverse event | Day 14 | Day 28 | Day 42 |
|---|---|---|---|
| Any adverse event | 97 (77.0%) | 46 (36.5%) | 70 (56.5%) |
| Pain at injection site | 85 (67.5%) | 17 (13.5%) | 49 (39.5%) |
| Fatigue | 50 (39.7%) | 26 (20.6%) | 44 (35.5%) |
| Headache | 32 (25.4%) | 22 (17.5%) | 23 (18.5%) |
| Fever | 10 (7.9%) | 3 (2.4%) | 7 (5.6%) |
| Local redness | 5 (4.0%) | 1 (0.8%) | 2 (1.6%) |
| Vomiting | 8 (6.3%) | 6 (4.8%) | 6 (4.8%) |
| Diarrhea | 13 (10.3%) | 11 (8.7%) | 9 (7.3%) |
| Muscle pain | 19 (15.1%) | 11 (8.7%) | 17 (13.7%) |
| Joint pain | 22 (17.5%) | 11 (8.7%) | 17 (13.7%) |
| Insomnia | 17 (13.5%) | 11 (8.7%) | 10 (8.1%) |

Variables are presented as n (%)

**Table S2. Baseline predictors of day 42 anti-SARS-CoV-2 RBD IgG titers according to linear regression model.** SLE activity is measured with SLEDAI.

| | **β** | **95% CI** | **p-value** |
|---|---|---|---|
| Age, years | -0.47 | -1.8, 0.88 | 0.5 |
| Male sex | -55 | -121, 10 | 0.10 |
| SLEDAI 2K >4 on day 1 | -19 | -81, 42 | 0.5 |
| C3, g/L | 43 | -70, 156 | 0.4 |
| dsDNA antibodies, IU/mL | 0.04 | -0.05, 0.14 | 0.4 |
| IFN | -2.9 | -6.9, 1.1 | 0.2 |
| IgA, g/L | 0.62 | -14, 15 | >0.9 |
| IgG, g/L | 2.0 | 0.35, 3.6 | 0.018 |
| IgM, g/L | 12 | -0.66, 25 | 0.063 |
| Lymphocytes, day 1 (G/L) | 9.0 | -29, 47 | 0.6 |
| Corticosteroids low | -18 | -64, 28 | 0.4 |
| Corticosteroids high | -51 | -130, 28 | 0.2 |
| Hydroxychloroquine | -22 | -80, 36 | 0.5 |
| Azathioprine | -128 | -256, -0.22 | 0.050 |
| Belimumab | -15 | -89, 58 | 0.7 |
| Mycophenolate mofetil | -77 | -154 | 0.008 |
| Other immunosuppressor | 63 | -32, 157 | 0.2 |
| Methotrexate | -120 | -241 | <0.001 |

β: see Material and methods; CI = Confidence Interval; dsDNA: double stranded DNA; IFN: Interferon; RBD: Receptor Binding Domain; SLEDAI: SLE Disease Activity Index

**Table S3. Baseline B cell predictors of serum neutralizing activity at day 42 according to linear regression model**

| | β | 95% CI | p-value |
|---|---|---|---|
| Age, years | -0.01 | -0.02, 0.01 | 0.6 |
| Male sex | -0.76 | -1.7, 0.15 | 0.10 |
| At least one BILAG score ≥ B | -1.2 | -2.1, -0.27 | 0.011 |
| C3, g/L | 0.81 | -0.71, 2.3 | 0.3 |
| dsDNA antibodies, IU/mL | 0.00 | 0.00, 0.00 | 0.7 |
| Detectable IFN alpha | -0.06 | -0.11, 0.00 | 0.046 |
| Total serum IgA, g/L | -0.02 | -0.22, 0.17 | 0.8 |
| Total serum IgG, g/L | 0.01 | -0.01, 0.04 | 0.3 |
| Total serum IgM, g/L | 0.05 | -0.12, 0.23 | 0.5 |
| Corticosteroids ≤10mg/day | -0.04 | -0.67, 0.59 | >0.9 |
| Corticosteroids >10mg/day | -0.07 | -1.1, 1.0 | >0.9 |
| Hydroxychloroquine | -0.01 | -0.83, 0.80 | >0.9 |
| Azathioprine | -1.4 | -3.2, 0.30 | 0.10 |
| Belimumab | 0.26 | -0.75, 1.3 | 0.6 |
| Mycophenolate mofetil | -1.1 | -1.9, -0.34 | 0.005 |
| Methotrexate | -1.9 | -2.7, -1.0 | <0.001 |
| Other immunosuppressor | 0.26 | -1.0, 1.6 | 0.7 |

β: see Material and methods; BILAG: British Isles Lupus Assessment Group; CI = Confidence Interval; dsDNA: double stranded DNA; IFN: Interferon;

**Table S4: Results of linear regression model for baseline B cell subpopulations and serum neutralizing activity at day 42**

| | β* | 95% CI | p-value |
|---|---|---|---|
| Corticosteroids ≤10mg/day | -0.18 | -0.79, 0.43 | 0.6 |
| Corticosteroids >10mg/day | -1.0 | -2.1, 0.19 | 0.10 |
| Hydroxychloroquine | 0.05 | -0.90, 1.0 | >0.9 |
| Azathioprine | -0.85 | -2.8, 1.1 | 0.4 |
| Belimumab | 0.30 | -1.1, 1.7 | 0.7 |
| Mycophenolate mofetil | -1.6 | -2.6, -0.69 | 0.001 |
| Methotrexate | -1.4 | -2.4, -0.39 | 0.007 |
| Other immunosuppressor | 1.3 | -1.1, 3.7 | 0.3 |
| Marginal zone B lymphocytes, Day 1 (%) | -0.01 | -0.04, 0.02 | 0.5 |
| Autoreactive B lymphocytes, Day 1 (%) | -0.01 | -0.04, 0.03 | 0.7 |
| Naive B lymphocytes, Day 1 (%) | 0.04 | 0.01, 0.07 | 0.006 |
| Double negative B lymphocytes, Day 1 (%) | -0.01 | -0.08, 0.07 | 0.9 |
| Memory B lymphocytes, Day 1 (%) | -0.02 | -0.04, 0.00 | 0.10 |

β: see Material and methods; CI = Confidence Interval; *Log (D614G); Autoreactive B cells (CD21lowCD38low); Double Negative B cells (CD27-IgD-); Marginal Zone B cells (CD27+IgD+); Memory B cells (CD27+IgD-); Naïve B cells (CD27-IgD+). B cell subsets frequencies are measured in total B cells.

**Figure S1: Analysis of peripheral B cell subsets phenotyping**

Representative flow cytometry gating strategy for CD27+IgD- memory B cells (A), CD27+IgD+ Marginal Zone B cells (B), CD27-IgD- Double Negative B cells (C), CD27-IgD+ Naïve B cells (D), CD21lowCD38low B cells (E)

**Figure S2: Local and systemic reactions reported throughout the vaccination schedule.**
