## Supplementary material for "BNT162b2 vaccine-induced humoral and cellular responses against SARS-CoV-2 variants in Systemic Lupus Erythematosus": Figure S1

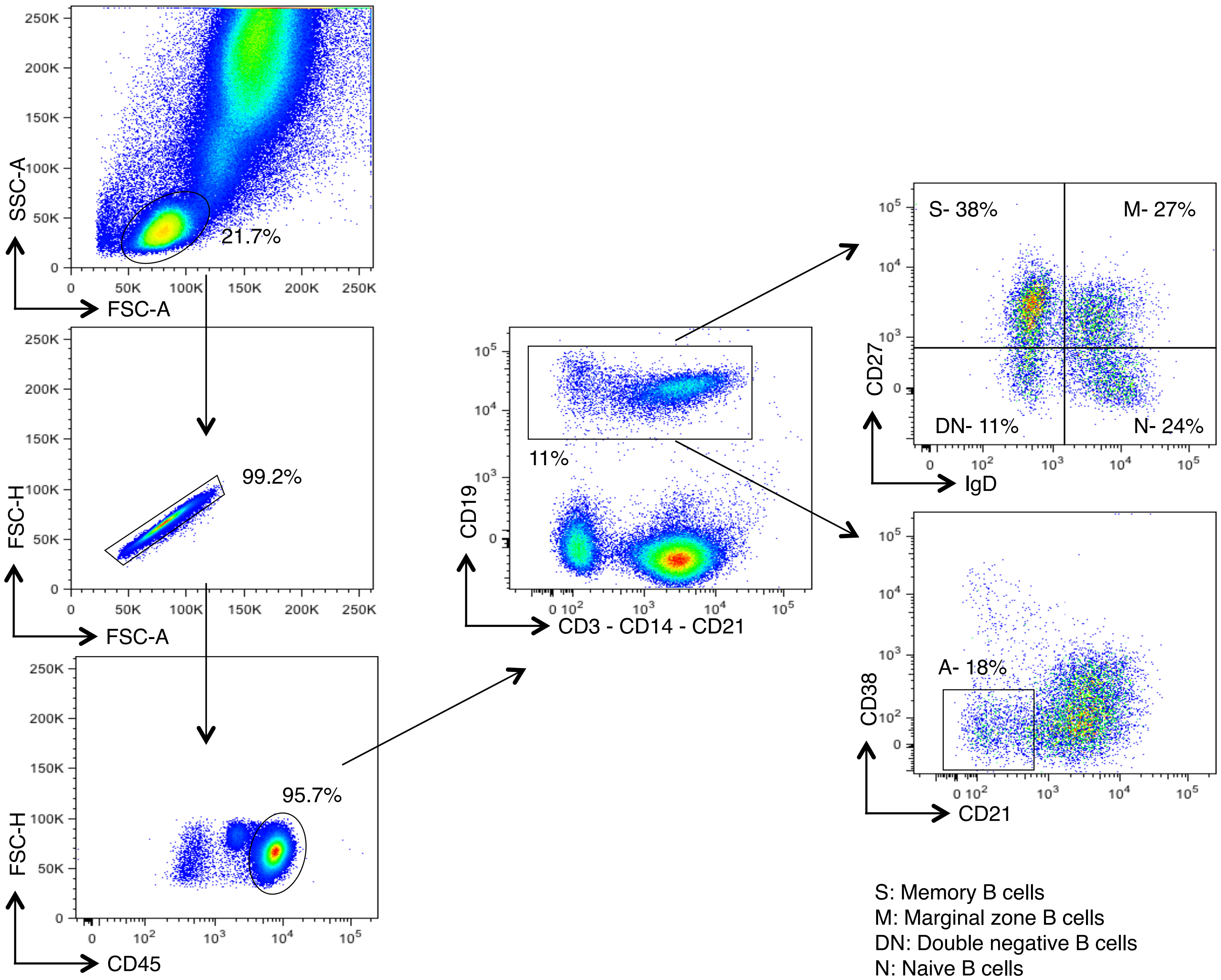


S: Memory B cells
M: Marginal zone B cells
DN: Double negative B cells
N: Naive B cells
A: CD21lowCD38low B cells
