## Supplementary figures and images for "BNT162b2 vaccine-induced humoral and cellular responses against SARS-CoV-2 variants in Systemic Lupus Erythematosus"

### Figure S2

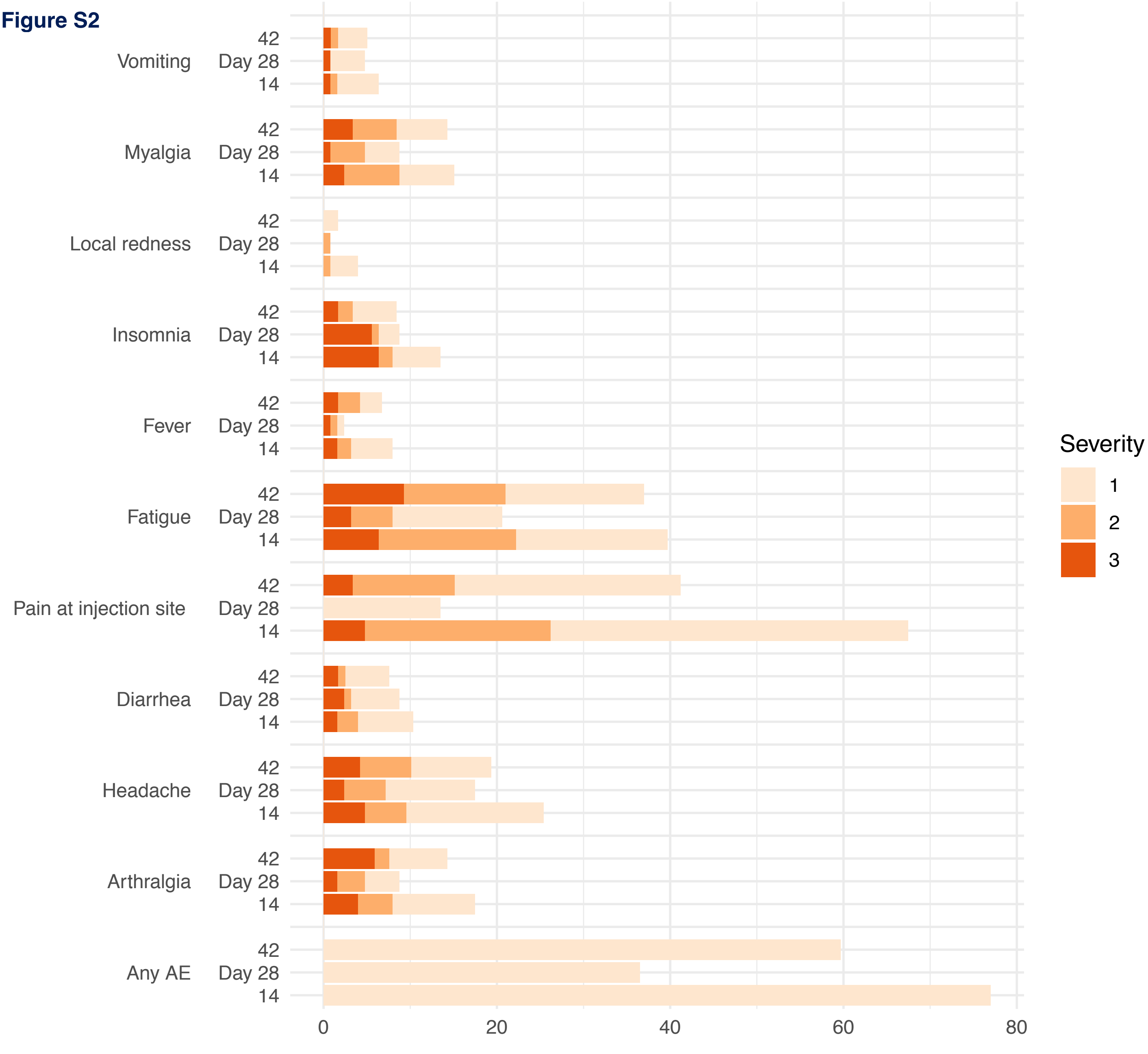
